# Dipstick-based pathogen detection for wastewater surveillance: Variability analysis using gage repeatability and reproducibility

**DOI:** 10.1101/2024.11.07.24316947

**Authors:** Shruti Ahuja, Avani Kulkarni, Kiran Kondabagil, Siddharth Tallur

## Abstract

COVID-19 redefined the outlook on pandemic preparedness, accelerating research toward establishing a global consortium for wastewater surveillance. Due to sample heterogeneity and low pathogen loads, microbial concentration remains a key challenge in developing low-cost, point-of-use wastewater monitoring assays. To address this challenge, we have developed a simplified version of dipstick method for RNA capture and isolation from sub-milliliter sample volumes, which simplifies RNA isolation. Given the manual steps involved in executing the dipstick method, variability is a major concern. In this work, we assessed dipstick variability through a multi-operator gage repeatability & reproducibility (gage R&R) study. We focused on detecting pepper mild mottle virus (PMMoV) and bacteriophage Phi6 in wastewater samples collected from a sewage pumping station at IIT Bombay. Our study demonstrated that the repeatability and reproducibility for the dipstick method are less than the acceptability limit of 30%, and could detect changes in PMMoV load associated with change in the population density due to summer break in our campus. Phi6 is a widely accepted surrogate for enveloped viruses such as SARS-CoV-2, and was therefore also chosen to demonstrate utility of this method for monitoring spread of infectious diseases. This work underscores the effectiveness of gage R&R in assessing and understanding the sources of variations in such assays, including operator-induced and part-to-part differences, essential for developing robust, manually-operated assays.

## I. INTRODUCTION

Wastewater surveillance provides real-time insights into spatio-temporal trends in spread of infectious diseases and collecting comprehensive epidemiological data for pandemic preparedness and rapid response [1, 2]. Characterized by its heterogeneous composition of biological materials including bodily fluids such as urine, feces, saliva, and respiratory and nasal secretions from asymptomatic and symptomatic individuals, wastewater poses significant challenges for pathogen monitoring [3–6]. Furthermore, factors such as rainfall, infection prevalence, and waste discharge contribute to data variability, complicating on-site wastewater surveillance efforts [7–9]. To simplify the workflow of wastewater surveillance, we have developed an instrument-free dipstick method for nucleic acid extraction from microbes in wastewater. Using this method, we have previously demonstrated concentration and isolation of nucleic acids from different microbes such as SARS-CoV-2, PMMoV, and Phi6 from wastewater samples with recovery efficiency comparable to commercially available kits [10]. This method includes thermal lysis of pathogens, followed by nucleic acid purification using paper dipsticks, leveraging the favorable nucleic acid binding and release kinetics of cellulose matrix [11, 12]. However, due to the manual nature of the method, it is important to understand the relative contributions of various sources of variation that impact assay variability.

Measurement system analysis (MSA) is one method of quantifying variability, that employs various statistical tools essential for examining statistical quality control [13]. It assesses the impact of measurement errors on variations in manufacturing processes and evaluates the accuracy, precision, and stability of a measurement system (MS) [14, 15]. MS errors can be systematic errors such as bias and non-linearity, or random errors associated with repeatability and reproducibility, often assessed through gage repeatability and reproducibility (gage R&R) studies. Gage R&R is a promising tool for assessing the accuracy, precision, and variability in biological assays, especially those involving complex and heterogeneous samples. Unlike traditional statistical methods such as ANOVA and t-tests, which primarily focus on comparing means and determining statistical significance between experimental conditions [16, 17], gage R&R provides a comprehensive analysis of variability by isolating and quantifying the effects of the measurement system, operator handling, and sample characteristics [18]. This method quantifies repeatability (intra-operator variation) and reproducibility (inter-operator variation), providing a clear understanding of reliability and ensuring consistent performance of the MS across different settings.

Gage R&R analysis has been recently utilized to study variability in a few biological assays. For instance, Betancourt-Rodriguez et al. employed gage R&R for validating a novel isothermal microcalorimetry method by analyzing the variability in heat flow measurements performed by two different operators [19]. Similarly, gage R&R has been applied to determine the total variation in a new digital technique measuring the volumetric healing process of free gingival grafts surrounding dental implants in ten patients [20]. Additionally, in cell engineering, gage R&R method has been used to validate a protocol to express and purify *α*-synuclein, with variability analyzed using descriptors from techniques like PAGE, IMA, IEF, HDX, and peptide mapping [21]. Gage R&R has also been applied to study the operator variability for instruments used in biological processes. For instance, gage R&R study has been used to study operator-to-operator variability amidst trained and in-training analysts for assessing the consistency of digital ECG systems and ECG mark placements at group levels [22]. Similarly, commercially available thermocyclers, such as Eco™ Real-Time PCR system (Illumina) have been validated by gage R&R studies. [23]. However, most of these gage R&R studies use the AIAG gage R&R approach developed by the Automotive Industry Action Group (AIAG), primarily used in the automotive industry and other manufacturing sectors [24]. AIAG gage R&R is suitable for manufacturing environments where the primary concern is the consistency and accuracy of measurement systems used for quality control and process optimization [25]. Evaluating the measurement process (EMP) gage R&R is an advanced version of R&R analysis, that incorporates more detailed analysis and modern statistical methods that can be applied to a wider range of industries, including biological and environmental studies [26]. EMP gage R&R provides a comprehensive analysis of variability, including interactions between different factors [27], which is crucial for understanding how various sources of variability affect the assay performance in biological contexts. It can accommodate different measurement designs and conditions, making it more versatile for various biological and environmental conditions as compared to other statistical tools [18, 27, 28].

In order to study the repeatability, reproducibility, accuracy and precision of the dipstick method to isolate and capture nucleic acids from 1 mL of wastewater sample, we have conducted a rigorous multi-operate EMP gage R&R analysis, as illustrated in Figure 1. The key contribution of this work are summarized below:

**FIG. 1.**
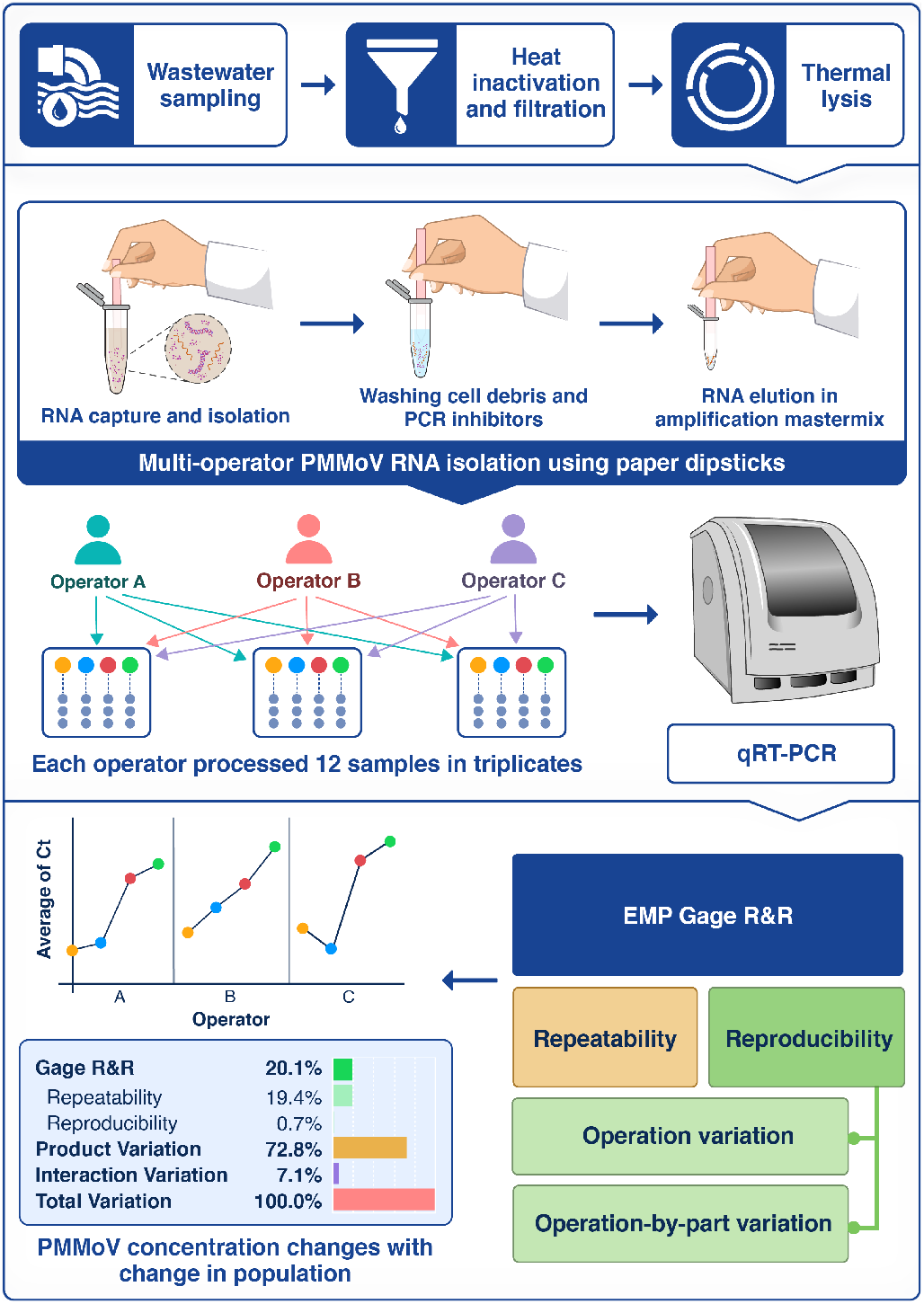
Schematic illustration of multi-operator gage R&R study for variability analysis of dipstick method employed in wastewater surveillance.

- The dipstick method was analyzed using both a simplified procedure with manual dipstick preparation and RNA elution, and a rigorous method with automated dipstick preparation and more elaborate procedure for RNA elution. These assay steps, which could have contributed to variability, were evaluated through EMP gage R&R. The gage R&R analysis for both methods confirmed that the simplified dipstick method did not negatively impact performance, validating overall reliability of the method.
- The variability of the simplified dipstick method was further evaluated through a multi-operator gage R&R study using wastewater samples from a sewage pumping station at IIT Bombay. The analysis showed that the dipstick method has gage R&R values below 30%, classifying it as a second-class monitor according to EMP guidelines, and indicating that the simplified dipstick method is within acceptable range of variability and reliable for field applications [29]. The utility for field applications was demonstrated by employing the method for detecting variations in PMMoV load associated with population density changes due to summer break on campus.

## II. MATERIALS AND METHODS

### A. Wastewater samples and pre-processing

The microbes and the concentration methods used in this study are outlined in Table I. The selected microbes include bacteriophage Phi6, commonly used as a surrogate for studying enveloped RNA viruses such as SARSCoV-2 [30], and pepper mild mottle virus (PMMoV), commonly utilized to assess fecal pollution in water samples and microbial water quality. PMMoV is abundantly present in human feces, making it a reliable control organism for normalizing pathogen loads in wastewater samples relative to population size [31]. Wastewater samples were collected following the sampling strategy outlined by the CDC [32], from the sewage pumping station at the IIT Bombay campus, Mumbai, India, encompassing both residential and academic zones.

**TABLE I.** Wastewater samples used in this study.

| Sample ID<sup>a</sup> | pH | Collection window | Dipstick method | Captured organism | Number of operators<sup>b</sup> | Purpose |
| --- | --- | --- | --- | --- | --- | --- |
| 1 | 7 | During break | Simplified & Rigorous | Phi6 | 2 (A,B) | Comparing simplified and rigorous dipstick method |
| 2 | 7 | Before break | Simplified | PMMoV, Phi6 | 3 (A,B,C) | Detecting change in PMMoV concentration due to summer break; variability analysis for simplified dipstick method |
| 3 | 8 |  |  |  |  |  |
| 4 | 7 | During break |  |  |  |  |
| 5 | 7 |  |  |  |  |  |

<sup>a</sup>Wastewater samples were collected from the sewage pumping station at IIT Bombay, Mumbai, India.
<sup>b</sup>Each operator performed measurements in triplicates.

This study utilized 5 wastewater samples, denoted by sample IDs 1 to 5. Wastewater sample ID 1 was collected to evaluate the performance variation between the simplified and rigorous dipstick method. Sample IDs 2 and 3 were collected before semester break, while sample IDs 4 and 5 were collected during semester break. Sample IDs 2 to 5 were used to study gage R&R using simplified dipstick method and for detecting changes in PMMoV concentrations associated with population fluctuations before and during the semester break. For sample IDs 2 to 5, bacteriophage Phi6 served as a process control.

A schematic of the sample pre-processing method is shown in supplementary information Figure S1. Briefly, 100 mL of raw grab wastewater samples were aseptically collected using sterile wide-mouth bottles (Tarsons, PP autoclavable). All the samples were collected between 9 am to 10 am in triplicates and stored at 4 °C before being processed within 24 hours of collection. The wastewater samples were heat inactivated at 60 °C for 30 min followed by dechlorination with 1 mL of 1 g*/*10mL of sodium thiosulfate [33, 34]. The pH levels were measured using pH indicator strips (MQuant®, Merck). For the dipstick assay, 1 mL of wastewater samples were spiked with bacteriophage Phi6 to attain a final concentration of 10^6^ PFU*/*mL (plaque forming units/milliliter). To mimic the natural wastewater conditions, the spiked wastewater samples were gently mixed using a shaker at 30 rpm for 10 min at room temperature. The samples were then filtered using a 5 μm mixed cellulose ester membrane filter (MF-Millipore) followed by a 1.6 μm glass fibre membrane filter to eliminate suspended particles.

### B. Dipstick preparation and method of use

Two different dipstick methods, labeled simplified and rigorous, were evaluated with distinct preparation techniques and usage protocols. Purification of nucleic acids released in 1 mL of the sample following thermal lysis at 60 °C was achieved in three steps as detailed in our previous work [10]:

- *Step 1* : Nucleic acid binding to the exposed region of the dipstick (i.e., portion of the dipstick that is not coated with wax)
- *Step 2* : Washing of exposed region of the dipstick to remove impurities
- *Step 3* : Elution of RNA in reverse transcription master-mix

The dipstick preparation was based on the methodology outlined by Mason & Botella (2020) [35]. For both methods, the dipsticks were prepared using Whatman grade 1 filter paper sheets (100 mm *×* 75 mm, GE Healthcare). Colored candle wax was melted at 80 °C on a hotplate until the wax was completely liquefied in a 120 mm borosilicate petri dish. The molten wax was infused into the cellulose filter paper, saturating a length up to 55 mm from one end to create a waterproof handle for holding the dipstick. Photographs of dipsticks prepared for simplified and rigorous methods are shown in supplementary information Figure S2. The subsequent preparation and usage for each method are described below:

- Simplified dipstick assay: For the simplified method, wax-infused cellulose sheets were manually cut using sterile scissors. The dipsticks were cut to a width of 2 mm and an overall length of 65 mm, including the wax-infused segment. The exposed section responsible for nucleic acid capture had a length of 10 mm, with the length of the triangular section at the tip being 1 mm. Initially, the cellulose dipstick was immersed in 1 mL of heat-lysed sample and dipped 10 to 15 times till it is completely soaked to facilitate nucleic acid capture. Subsequently, the dipstick was washed in 500 μL of wash buffer (10 mM Tris buffer adjusted to pH 8.0) by gently dipping it thrice to remove cellular debris and impurities. Finally, the dipstick was immersed in a 0.2 mL tube containing 20 μL reverse transcription master-mix and vigorously moved up and down 10 to 15 times.
- Rigorous dipstick assay: The rigorous dipstick method was developed to be more systematic, reduce manual handling, and allow for scalability in wastewater monitoring using dipsticks. For this method, wax-infused cellulose sheets were cut into 2 mm wide strips using a pasta maker as originally demonstrated by Mason et.al. [35] The steps for RNA isolation and purification from thermally lysed samples were similar to the simplified method, but with a few modifications. Initially, the dipstick was immersed in the sample for 45 s followed by washing in 500 μL of TE wash buffer thrice. The RNA captured on the dipstick was then eluted by immersing it for 60 s in 40 μL of reverse transcription master-mix.

The reverse transcription was performed using (iScript cDNA synthesis kit, Bio-Rad Laboratories) with 40 units of RNaseOUT™ recombinant ribonuclease inhibitor (Invitrogen). The steps involved in cDNA synthesis reaction are detailed in supplementary information Table S1.

### C. Real-time quantitative PCR (qPCR) for target amplification

To confirm the presence and quantify pepper mild mottle virus (PMMoV) and bacteriophage Phi6 in wastewater samples, real-time quantitative PCR (qPCR) was performed. For benchmarking the dipstick-based RNA extraction methods, RNA was also extracted from the wastewater samples using the Qiagen PowerWater® RNeasy® kit, according to the manufacturer’s instructions. Briefly, pre-processed samples were filtered through a 0.2 μm nylon filter (Axiva Sichem) using a vacuum filtration assembly, followed by mechanical bead beating for cell lysis. The purified RNA was eluted in 100 μL of RNase-free water, followed by cDNA synthesis. The cDNA generated from three methods (Qiagen kit-based RNA extraction, simplified dipstick assay, and rigorous dipstick assay) served as templates for qPCR amplification. No template control (NTC) was included in each qPCR run. All qPCR reactions were performed in triplicates. The qPCR reaction was performed using Brilliant III Ultra-Fast SYBR® Green QPCR Master Mix (Agilent Technologies) in Agilent technologies Stratagene MX3000P for 40 cycles. Details of the primers for PMMoV and bacteriophage Phi6, as well as the qPCR thermal cycling conditions are detailed in Table S2, and S3 in supplementary material.

### D. Multi-operator study and gage R&R variability analysis

To evaluate the accuracy and precision of the dipstick method in pathogen detection from wastewater samples, we conducted a multi-operator study combined with a gage repeatability and reproducibility (Gage R&R) analysis using the EMP (Evaluating the Measurement Process) approach. While traditional gage R&R studies assess repeatability and reproducibility, EMP goes further by focusing on a system’s overall performance and its suitability for reliably measuring a specific process or output [36, 37]. This analysis was performed using the JMP® statistical software, focusing on variability across operators, samples, and interactions. The multi-operator study was conducted to:

- Compare the variability between the simplified and rigorous dipstick methods: Two operators (A and B) operated both the dipstick assays in triplicates on wastewater sample ID 1 spiked with 10^6^ PFU*/*mL of bacteriophage Phi6
- Assess operator variability and field applicability for the simplified dipstick method: Three operators (A, B, and C) analyzed wastewater samples ID 2 to 5 collected before and during the semester break to assess repeatability and reproducibility, and evaluate reliability for detecting change in PMMoV load as compared to process control

We performed a crossed gage R&R study, wherein each operator performed the dipstick method on each sample in triplicates under similar conditions. This crossed approach completely overlaps each factor, such as operator and sample, allowing for comprehensive variability analysis across these interactions [38, 39]. The input parameters included *part* (wastewater sample with unknown concentrations of PMMoV), *operator* (technicians A, B and C), and *measurement* (threshold cycle (Ct) value obtained through RT-qPCR serving as the primary quantitative measure for gage R&R analysis). Additionally, the sigma multiplier was set to 6 standard deviations (*±*3 sigma), representing the process width. This multiplier is standard in EMP studies to capture a broad range of natural process variation in assay performance [37].

Residual Maximum Likelihood (REML) was applied to estimate the variance components, as it effectively manages unbalanced data and offers robust variance component estimates in biological assays [40]. The variance components assessed included repeatability, reproducibility, and interaction variation [41]. Repeatability (intraoperator variability) or equipment variation corresponds to the *within-assay* variation in the variance components. A measurement is considered repeatable if a single operator consistently obtains similar results when measuring the same sample multiple times. Reproducibility (interoperator variation) or appraiser variability is analogous to *between-operator* variability in the variance components and measures precision across different operators. If multiple operators achieve consistent results using the same dipstick method on the same sample, the measurement is considered reproducible. Additionally, we evaluated product variation (or sample variation), which explains the variability due to factors like dipstick manufacturing, sample heterogeneity, and differences in viral load across samples. Operator-sample interaction or interaction variation, which accounts for the variability introduced by the operator and sample combination, was also assessed.

Initially, operators A and B assessed the variability in simplified and rigorous dipstick method. Once it was established that the variation in both the methods was comparable, a more thorough study was conducted with three operators A, B and C to study the simplified dipstick method to track change in PMMoV load with change in population. The two-operator analysis for sample ID 1 included 36 measurements (1 wastewater sample processed in triplicate) with two operators testing both dipstick methods, and Ct measurements further obtained in triplicates. Similarly, the gage R&R study for sample IDs 2 to 5 to assess operator-to-operator variability involved 108 measurements (4 wastewater samples processed in triplicates), each tested three times by three different operators, and Ct measurements further obtained in triplicates. Details of the samples are presented in Table I.

## III. RESULTS AND DISCUSSION

### A. Simplified dipstick method performs at par with the rigorous dipstick method

Variability analysis was performed for the isolation and purification of bacteriophage Phi6 spiked in 1 mL wastewater (sample ID 1), to attain a final concentration of 10^6^ PFU*/*mL. As shown in Figure 2(a), the average Ct values for sample ID 1 collected and processed in triplicates by operators A and B were consistent with a grand mean of 25.14. The variance components analysis as shown in supplementary information Figure S3 revealed that there is negligible variation due to the method or operator handling. Similarly, the operator-method interaction was minimal, confirming there was negligible operator and method dependency. The total observed variation was primarily due to repeatability. These results agree with the EMP gage R&R analysis shown in supplementary information Figure S3, which demonstrated minimal operator-to-operator variability. The product variation (which accounts for variation due to the dipstick manufacturing) and interaction variation (variation due to operator-sample combination) were negligible.

**FIG. 2.**
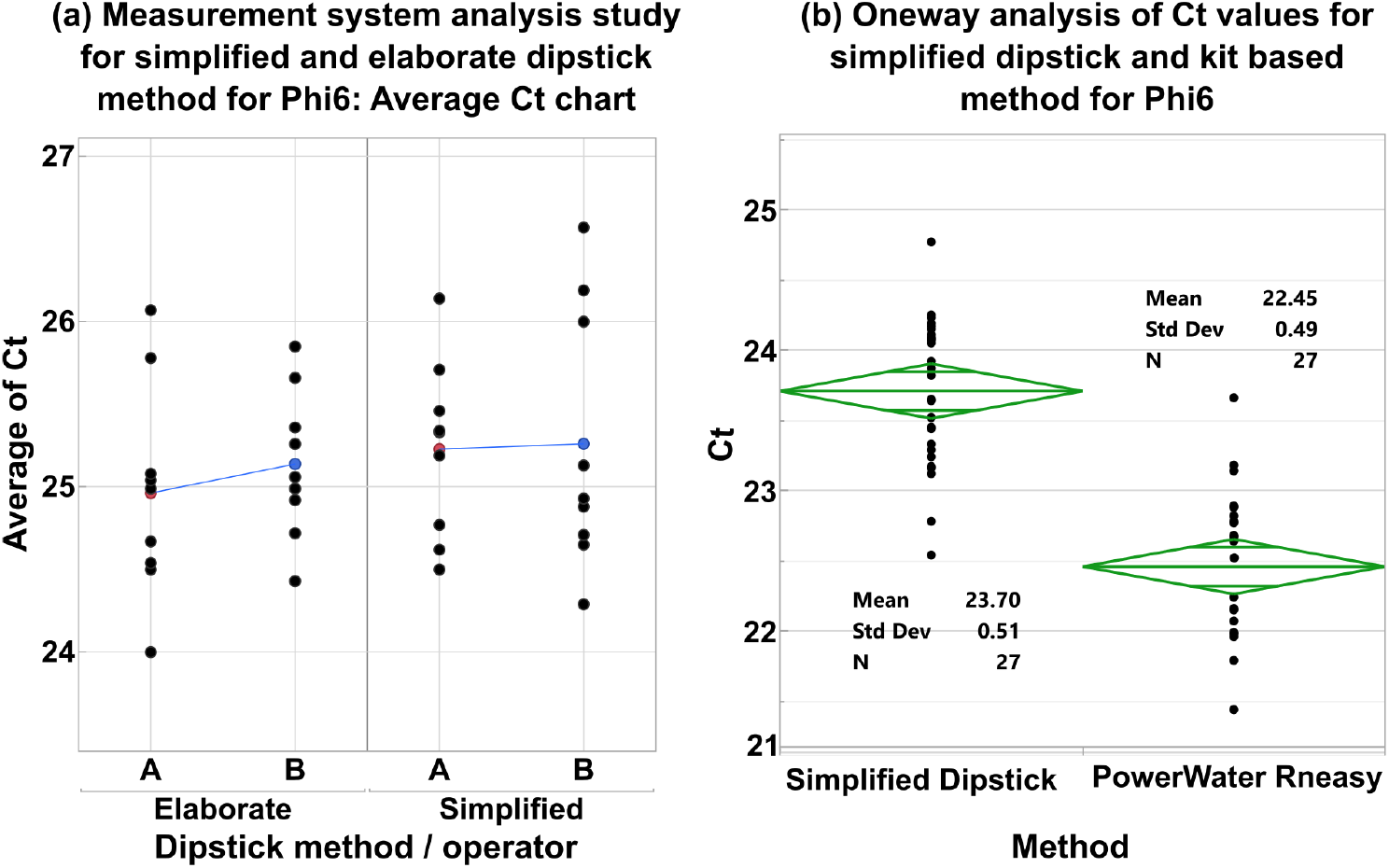
(a) Comparison of average Ct values for bacteriophage Phi6 RNA captured using the rigorous and simplified dipstick method by operators A and B in sample ID 1. (b) One-way analysis of Ct values for bacteriophage Phi6 across samples 2 to 4, comparing the simplified dipstick method with the Qiagen PowerWater® RNeasy® kit method.

Following the comparison between the simplified and rigorous dipstick methods, we evaluated the performance of the simplified dipstick method against the Qiagen PowerWater® RNeasy® kit for detecting bacteriophage Phi6 in samples IDs 2 to 4. The one-way analysis mean diamond plots in Figure 2(b), indicate that the spread in Ct values of both simplified dipstick and commercial kit based method are comparable. Additionally, Figure 2(b) and the distribution chart in supplementary information Figure S4 show that the mean Ct value for the kit-based method was 22.45 with a standard deviation of 0.49 while the simplified dipstick method had a mean Ct value of 23.70 with a comparable standard deviation of 0.51.

### B. Simplified dipstick method has acceptable variability to detect changes in viral load in wastewater samples

To further study variability sources in the simplified dipstick method, we conducted a multi-operator study to detect variations in PMMoV concentration corresponding to population change due to summer break on IIT Bombay campus. As shown in Figure 3, supported by Figure 4(a), the average Ct values for each operator increased in samples collected during the semester break (sample IDs 4 and 5) as compared to those collected before the break (sample IDs 2 and 3), indicating a decrease in PMMoV concentration during summer break. Additionally, as shown in Figure 4(a), the mean average of Ct by operator, and the Ct value spread for all operators was consistent (approximately 33) across all the samples. These observations were further corroborated by the EMP gage R&R results shown in Figure 3(a), where reproducibility (operator-to-operator variation) was remarkably low at just 0.7% followed by operator-sample ID interaction that contributed merely 7.1%. The largest contributor to variability was product variation, which accounted for 72.8%. Repeatability, which reflects intraoperator variation, contributed 19.4%.

**FIG. 3.**
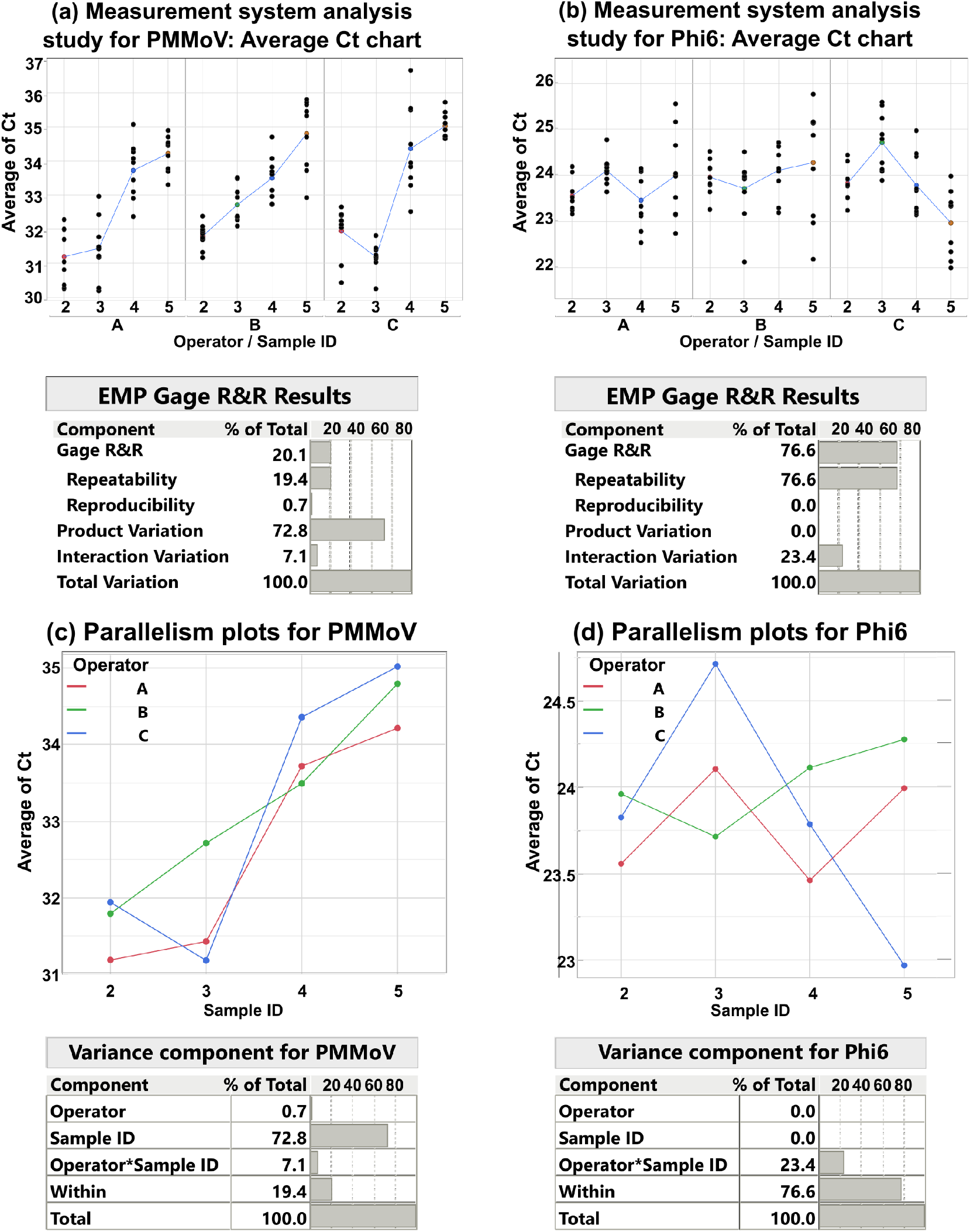
(a) Average Ct values for sample IDs 2 to 5, processed by operators A, B, and C, along with EMP gage R&R results for (a) PMMoV, and (b) bacteriophage Phi6. Parallelism plots illustrating change in average Ct for sample IDs 2 to 5 processed using simplified dipstick method by operators A, B, and C for (c) PMMoV, and (d) bacteriophage Phi6.

**FIG. 4.**
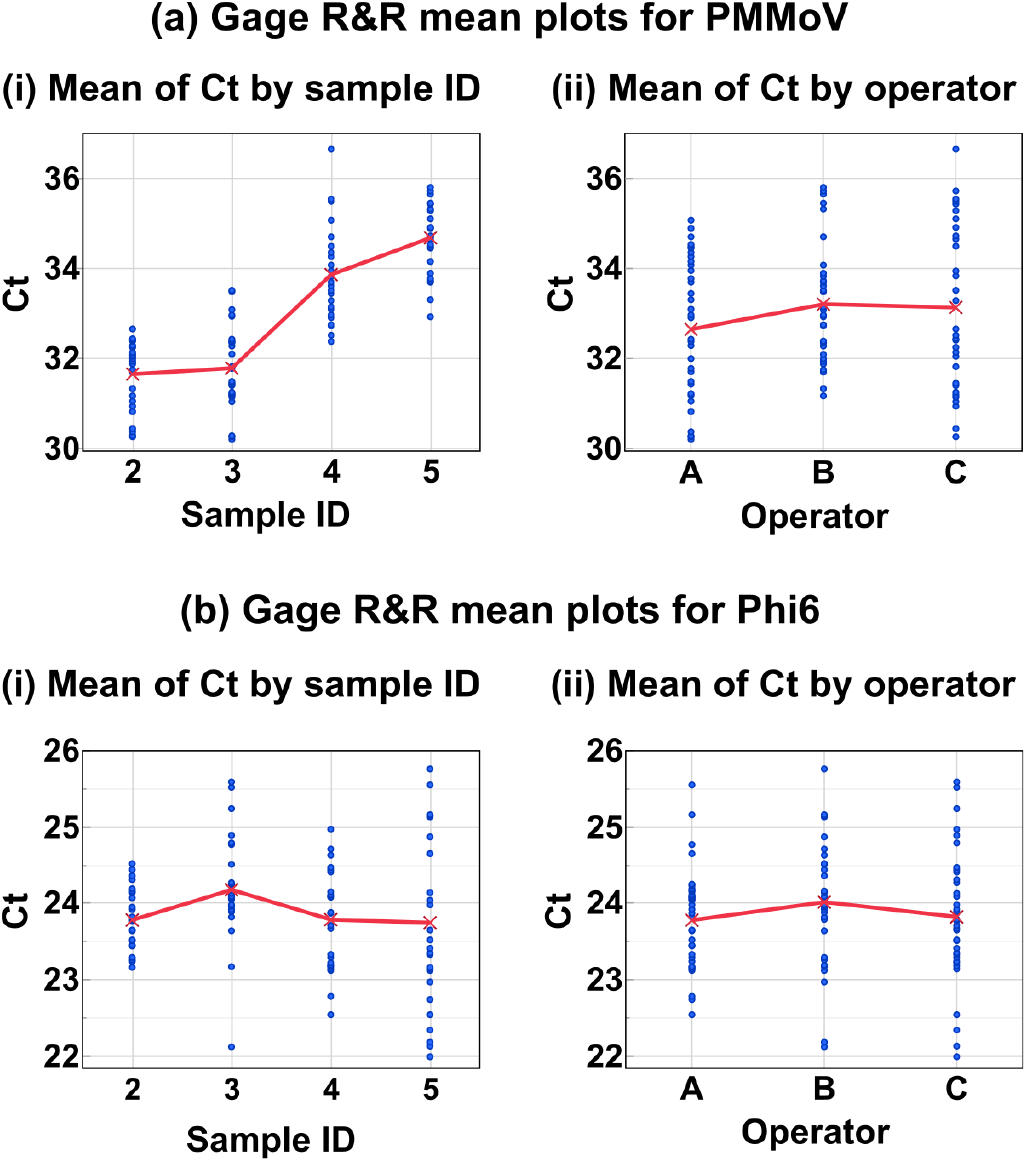
(a) Gage R&R mean plots for simplified dipstick method to detect PMMoV in sample IDs 2 to 5, illustrating variation of Ct with various parameters: (i) Ct vs. sample ID, and (ii) Ct vs. operator. (b) Gage R&R mean plots for simplified dipstick method to detect bacteriophage Phi6 in sample IDs 2 to 5, illustrating variation of Ct with various parameters: (i) Ct vs. sample ID, and (ii) Ct vs. operator.

The variance components analysis shown in Figure 3(c) was in agreement with the EMP gage R&R results, with minimal operator variability contributing only 0.7% and the majority of the variability arising from sample ID (72.8%) and operator-sample ID interaction variation accounting for only 7.1%. The total gage R&R was thus 20.1%, classifying it as a second-class monitor according to EMP guidelines, indicating that the simplified dipstick method is within acceptable range of variability and reliable for field applications [29].

For bacteriophage Phi6 (used as process control), the average Ct values were similar across different samples and operators (approximately 24) as shown in Figure 3(b), Figure 4(b), Similarly, as illustrated in Figure 4(b), both the mean Ct values for each operator and the Ct value spread were consistent across all samples. These observations agree with the EMP gage R&R results as shown in Figure 3(b), where both reproducibility and product variation were minimal, indicating negligible variability due to the operators or the dipstick method itself. However, the interaction variation was relatively higher for Phi6 at 23.4% compared to 7.1% for PMMoV. The primary source of variability for Phi6 detection was repeatability (accounting for 76.6%). The variance components as shown in Figure 3(d), mirrored the EMP gage R&R results.

## C. Discussion

The simplified dipstick method involves manual handling, which could introduce operator-to-operator variation, particularly during the elution step. To determine if such variation is significant, we compared the RT-qPCR Ct values as a readout by using Phi6 RNA isolated by simplified and rigorous dipstick method as a template to assess variance components through EMP gage R&R analysis. The analysis assessed variability due to the dipstick method, operator, and operator-method interaction, as well as the repeatability and reproducibility of the assays. While comparing the simplified and rigorous dipstick methods, the minimal operator-to-operator variability confirms that both the methods produced consistent results across operators. Furthermore, the negligible part-to-part (sample-to-sample) variation was in agreement with the experimental conditions, as the spiked bacteriophage Phi6 concentration was the same across all the samples. The gage R&R results confirmed that most of the observed variability was due to repeatability, and the mean average Ct for both the dipstick methods was similar. Both the methods were highly consistent in performance, and the simplified dipstick method performed at par with the rigorous dipstick method, with no significant performance differences, yielding reproducible results across operators. The minimal product and interaction variation further demonstrated that there was no significant variation in the dipstick method or in the interaction between operators and the dipstick methods. Additionally, we compared the kit-based method with simplified dipstick method. It was observed that even though the mean Ct values differ due to difference in recovery efficiency, the standard deviations for both methods were similar. This indicates comparable variability in Ct values between the two methods, promising the suitability of simplified dipstick method for field applications.

Field applicability of simplified dipstick method was further studied by three operators to detect changes in PMMoV load with change in population. All the three operators A, B and C successfully detected PMMoV concentration declining (i.e., increase in Ct value) in samples collected during semester break. The minimal spread in Ct values for the three operators while detecting PMMoV and the process control bacteriophage Phi6 as shown in Figure 4(a) and (b), emphasizes minimal operator-tooperator variability. For bacteriophage Phi6, the consistent Ct spread across the samples was expected, as the spiked concentration of bacteriophage Phi6 concentration was the same across all samples. The low variability in operator-sample ID interaction for PMMoV further demonstrated minimal influence of specific operator handling on sample variability. However, the interaction variation for Phi6 was comparatively higher, largely caused by the handling of operator C as demonstrated in the parallelism plots shown in Figure 3(d), which show a slight deviation in Ct values for sample IDs 3 and 5. Repeatability variation for Phi6 reflects intra-operator variation due to manufacturing variations in dipstick, and the different steps involved in executing the assay including washing and elution. This suggests room for further optimization of the method and dipstick design.

The primary source of variability for PMMoV detection observed in product variation was due to the natural fluctuations in PMMoV concentration due to change in population across the different samples collected over time. This high product variation is expected, given the dynamic nature of wastewater viral loads as population density and activity change. The total gage R&R at less than 30% indicates acceptable variability [27], demonstrating the assay’s reliability in consistently detecting target pathogens, even with moderate operator and sample variation, suitable for field applications.

## IV. CONCLUSION AND FUTURE WORK

This study demonstrates the utility of gage R&R analysis as a robust method to validate a biological assay when applied to complex and heterogeneous environmental samples such as wastewater. By systematically isolating and quantifying variations due to the measurement system, operator handling, and the samples themselves, gage R&R allows for a detailed assessment of both repeatability and reproducibility. The gage R&R analysis suggested that the variability was primarily attributed to within-sample repeatability, followed by operator-sample interaction and negligible due to operator-to-operator handling, indicating that the dipstick method consistently produced stable results across different operators and samples. This highlights the robustness of the simplified dipstick method. Unlike ANOVA and t-tests, which are primarily used to determine statistical significance between experimental conditions, gage R&R provides a nuanced understanding of how both the measurement system and human factors influence assay performance, making it indispensable for quality control and method validation in biological research. This shows that gage R&R extends beyond just the dipstick method evaluated here.

Our work shows that it can be widely applied in laboratory assays and field-deployed diagnostic methods to ensure the reproducibility of results across different settings, operators, and batches. This is particularly relevant when developing products or assays that will be used in decentralized or resource-constrained environments, where manual methods and operator variability may have a larger impact. The acceptable variability in the simplified dipstick method to isolate and purify nucleic acid highlight the field applicability of the test. With up-scaled manufacturing the dipstick method can be integrated with a field-portable RT-qPCR devices, or rapid detection techniques like loop-mediated isothermal amplification (LAMP), and a lateral flow assay to ensure on-site wastewater monitoring, making the setup more accessible in low-resource or remote areas.

## Supporting information

Supplementary information with additional figures and tables.

Supplementary video overview of the manuscript.

## DATA AVAILABILITY

The data that support the findings of this study are available upon reasonable request from the authors.

## ACKNOWLEDGMENT

S.A. acknowledges the Prime Minister’s Research Fellows (PMRF) Scheme for supporting her Ph.D. fellowship. This work was partially supported by grants from Department of Science & Technology (DST), Ministry of Science & Technology, Government of India [grant no. DST/IC/IC-IMPACTS/2022/P-13] and Wadhwani Research Centre for Bioengineering (WRCB) at IIT Bombay [grant DO/2022-WRCB002-076]. The authors thank Ms. Rutuja Chalke at IIT Bombay for help with preparing Figure 1 for this manuscript.

## References

[1] W.-y. Ng, W. Thoe, R. Yang, W.-p. Cheung, C.-k. Chen, K.-h. To, K.-m. Pak, H.-w. Leung, W.-k. Lai, T.-k. Wong, et al., The city-wide full-scale interactive application of sewage surveillance programme for assisting real-time covid-19 pandemic control–a case study in hong kong, Science of The Total Environment 875, 162661 (2023).

[2] K. Mao, K. Zhang, W. Du, W. Ali, X. Feng, and H. Zhang, The potential of wastewater-based epidemiology as surveillance and early warning of infectious disease outbreaks, Current Opinion in Environmental Science & Health 17, 1 (2020).

[3] A. Tiwari, S. Adhikari, D. Kaya, M. A. Islam, B. Malla, S. P. Sherchan, A. I. Al-Mustapha, M. Kumar, S. Aggarwal, P. Bhattacharya, et al., Monkeypox outbreak: Wastewater and environmental surveillance perspective, Science of the total environment 856, 159166 (2023).

[4] A. Tiwari, E. Radu, N. Kreuzinger, W. Ahmed, and T. Pitkänen, Key considerations for pathogen surveillance in wastewater, Science of The Total Environment, 173862 (2024).

[5] X. Li, J. Kulandaivelu, Y. Guo, S. Zhang, J. Shi, J. O’Brien, S. Arora, M. Kumar, S. P. Sherchan, R. Honda, et al., Sars-cov-2 shedding sources in wastewater and implications for wastewater-based epidemiology, Journal of hazardous materials 432, 128667 (2022).

[6] K. Bibby, A. Bivins, Z. Wu, and D. North, Making waves: plausible lead time for wastewater based epidemiology as an early warning system for covid-19, Water Research 202, 117438 (2021).

[7] L. Pillay, I. D. Amoah, S. Kumari, and F. Bux, Potential and challenges encountered in the application of wastewater-based epidemiology as an early warning system for covid-19 infections in south africa, Acs Es&T Water 2, 2105 (2022).

[8] J. Black, P. Aung, M. Nolan, E. Roney, R. Poon, D. Hennessy, N. D. Crosbie, D. Deere, A. R. Jex, N. John, et al., Epidemiological evaluation of sewage surveillance as a tool to detect the presence of covid-19 cases in a low case load setting, Science of The Total Environment 786, 147469 (2021).

[9] J. Saththasivam, S. S. El-Malah, T. A. Gomez, K. A. Jabbar, R. Remanan, A. K. Krishnankutty, O. Ogunbiyi, K. Rasool, S. Ashhab, S. Rashkeev, et al., Covid-19 (sars-cov-2) outbreak monitoring using wastewater-based epidemiology in qatar, Science of the Total Environment 774, 145608 (2021).

[10] S. Ahuja, S. Tallur, and K. Kondabagil, Simultaneous microbial capture and nucleic acid extraction from wastewater with minimal pre-processing and high recovery efficiency, Science of The Total Environment 918, 170347 (2024).

[11] Y. Zou, M. G. Mason, Y. Wang, E. Wee, C. Turni, P. J. Blackall, M. Trau, and J. R. Botella, Nucleic acid purification from plants, animals and microbes in under 30 seconds, PLoS biology 15, e2003916 (2017).

[12] B. Boese, K. Corbino, and R. Breaker, In vitro selection and characterization of cellulose-binding rna aptamers using isothermal amplification, Nucleosides, Nucleotides and Nucleic Acids 27, 949 (2008).

[13] R. Peruchi, A. d. Paiva, P. Balestrassi, J. Ferreira, and R. Sawhney, Weighted approach for multivariate analysis of variance in measurement system analysis, Precision Engineering 38, 651 (2014).

[14] S. S. Kamble, A. Gunasekaran, A. Ghadge, and R. Raut, A performance measurement system for industry 4.0 enabled smart manufacturing system in smmes-a review and empirical investigation, International journal of production economics 229, 107853 (2020).

[15] M. M. Wu, S.-T. Chan, D. Mazumder, D. Tamborini, K. A. Stephens, B. Deng, P. Farzam, J. Y. Chu, M. A. Franceschini, J. Z. Qu, et al., Improved accuracy of cerebral blood flow quantification in the presence of systemic physiology cross-talk using multi-layer monte carlo modeling, Neurophotonics 8, 015001 (2021).

[16] W. C. Driscoll, Robustness of the anova and tukey-kramer statistical tests, Computers & Industrial Engineering 31, 265 (1996).

[17] C. Bertinetto, J. Engel, and J. Jansen, Anova simultaneous component analysis: A tutorial review, Analytica Chimica Acta: X 6, 100061 (2020).

[18] D. J. Wheeler, An honest gauge r&r study, Manuscript (2009).

[19] J. Betancourt-Rodríguez, V. M. Zamora-Gasga, J. A. Ragazzo-Sánchez, J. A. N. Zapata, and M. Calderón-Santoyo, A standardized method for genus colletotrichum characterization by isothermal microcalorimetry using thermokinetic parameters, Journal of Microbiological Methods 204, 106651 (2023).

[20] C. Docampo-Vázquez, T. Gragera-Alia, M. Fernández-Domínguez, Á. Zubizarreta-Macho, and J. M. Aragoneses-Lamas, Novel digital technique for measuring the volumetric healing process of free gingival grafts surrounding dental implants, Frontiers in Dental Medicine 5, 1372312 (2024).

[21] H. Thakkar, S. Chatterjee, P. Saxena, R. Eerla, S. Wagh, A. Khairnar, and R. P. Shah, Cell-engineered recombinant α-synuclein: A gage r&r validated protocol, Journal of Proteome Research 23, 16 (2023).

[22] R. A. Ross, C. M. Foley, H. M. Jones, and M. A. Osinski, A method for assessing and monitoring consistency of nonclinical ecg analysis, Journal of Pharmacological and Toxicological Methods 116, 107189 (2022).

[23] Illumina, Eco™ thermal and optical systems deliver high precision (Technical Note: Real-Time PCR), https://www.illumina.com/documents/products/technotes/technote_eco_uniformity.pdf (2024), accessed: October 2024.

[24] C. LLC, Ford motor company, general motors corporation, Potential Failure Mode and Effect Analysis (FMEA), 67 (2008).

[25] E. Bottani, R. Montanari, A. Volpi, and L. Tebaldi, Statistical process control of assembly lines in manufacturing, Journal of Industrial Information Integration 32, 100435 (2023).

[26] D. J. Wheeler and R. W. Lyday, Evaluating the measurement process (SPC Press, Incorporated, 1989).

[27] D. J. Wheeler, EMP III using imperfect data (SPS Press, 2006).

[28] D. J. Wheeler, Range based analysis of means (SPC Press, 2003).

[29] D. J. Wheeler, EMP III (Evaluating the Measurement Process): Using Imperfect Data (SPC PRESS (Statistical Process Control), 2006).

[30] S. Torii, H. Furumai, and H. Katayama, Applicability of polyethylene glycol precipitation followed by acid guanidinium thiocyanate-phenol-chloroform extraction for the detection of sars-cov-2 rna from municipal wastewater, Science of The Total Environment 756, 143067 (2021).

[31] S.-Y. Hsu, M. Bayati, C. Li, H.-Y. Hsieh, A. Belenchia, J. Klutts, S. A. Zemmer, M. Reynolds, E. Semkiw, H.-Y. Johnson, T. Foley, C. G. Wieberg, J. Wenzel, M. C. Johnson, and C.-H. Lin, Biomarkers selection for population normalization in sars-cov-2 wastewater-based epidemiology, Water research 223, 118985 (2022).

[32] CDC, Developing a wastewater surveillance sampling strategy (Centers for Disease Control and Prevention, 2020).

[33] M. Lau, P. Monis, G. Ryan, A. Salveson, N. Fontaine, J. Blackbeard, S. Gray, and P. Sanciolo, Selection of surrogate pathogens and process indicator organisms for pasteurisation of municipal wastewater—a survey of literature data on heat inactivation of pathogens, Process Safety and Environmental Protection 133, 301 (2020).

[34] K. M. Babler, A. Amirali, M. E. Sharkey, S. L. Williams, M. M. Boone, G. A. Cosculluela, B. B. Currall, G. S. Grills, J. Laine, C. E. Mason, et al., Comparison of electronegative filtration to magnetic bead-based concentration and v2g-qpcr to rt-qpcr for quantifying viral sars-cov-2 rna from wastewater, ACS ES&T Water 2, 2004 (2022).

[35] M. G. Mason and J. R. Botella, Rapid (30-second), equipment-free purification of nucleic acids using easy-to-make dipsticks, Nature Protocols 15, 3663 (2020).

[36] B. S. Ramirez and J. G. Ramírez, Douglas Montgomery’s Introduction to Statistical Quality Control: A JMP Companion (Sas Institute, 2018).

[37] M. A. Durivage, Practical attribute and variable measurement systems analysis (MSA): a guide for conducting Gage R&R studies and test method validations (Quality Press, 2015).

[38] F. d. Almeida, T. De Paula, R. Leite, G. Gomes, J. Gomes, A. Paiva, and P. Balestrassi, A multivariate gr&r approach to variability evaluation of measuring instruments in resistance spot welding process, Journal of Manufacturing Processes 36, 465 (2018).

[39] R. A. M. Marques, R. B. D. Pereira, R. S. Peruchi, L. C. Brandão, J. R. Ferreira, and J. P. Davim, Multivariate gr&r through factor analysis, Measurement 151, 107107 (2020).

[40] R. Rameez, S. Jahageerdar, J. Jayaraman, T. I. Chanu, R. Bangera, and A. Gilmour, Evaluation of alternative methods for estimating the precision of reml-based estimates of variance components and heritability, Heredity 128, 197 (2022).

[41] R. S. Kenett and G. Shmueli, Clarifying the terminology that describes scientific reproducibility, Nature methods 12, 699 (2015).

