## Supplementary information with additional figures and tables. for "Dipstick-based pathogen detection for wastewater surveillance: Variability analysis using gage repeatability and reproducibility"

### 1 Wastewater sample pre-processing

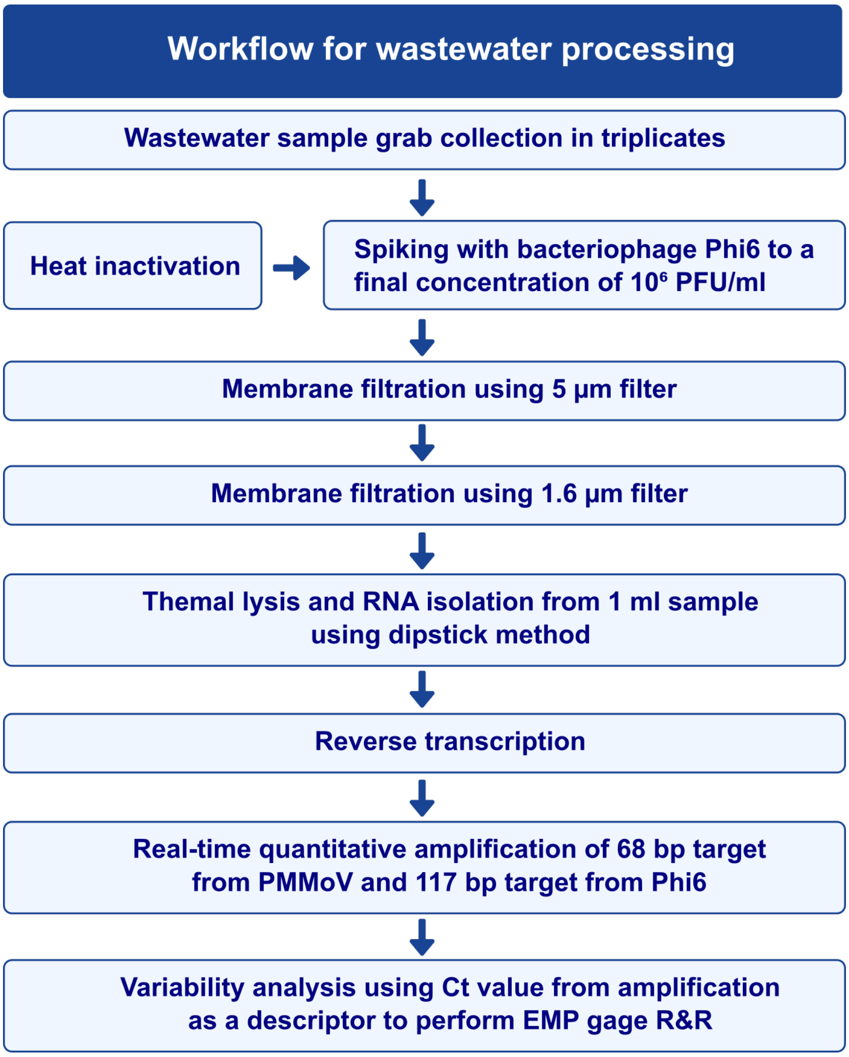


Figure. S1: Schematic representation of wastewater sample processing and variability analysis workflow for dipstick method.

#### 2 Dipstick preparation

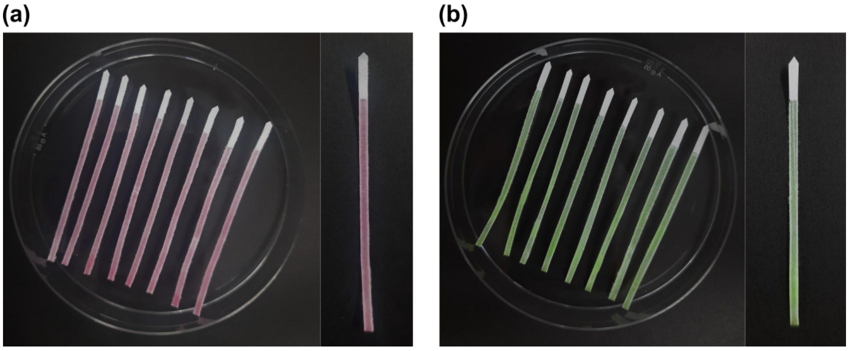


Figure. S2: Photograph of dipsticks prepared: (a) using pasta maker for rigorous dipstick method; (b) manual cutting for simplified dipstick method.

#### 3 Real-time quantitative PCR (qPCR) conditions

Table S1: Reverse transcription conditions

| Step | Temperature | Time (minutes) |
|---|---|---|
| Priming | 25 °C | 5 |
| Reverse transcription | 46 °C | 20 |
| Enzyme inactivation | 95 °C | 1 |

Table S2: PCR primers used in this study

| Microbe | Target gene | Forward primer (5′ – 3′) | Reverse primer (5′ – 3′) | Size (bp) |
|---|---|---|---|---|
| Bacteriophage Phi6 | M segment | GAATCATATGCGCTACC AAGGCATCAAC | CATAGAATTCTGGG AGGAGCAGCG-GAGA | 117 |
| PMMoV | 1878 bp to 1901 bp; 1945 bp to 1926 bp[a] | GAGTGGTTTGACCTT AACGTTTGA | TTGTCGGTTGC AATGCAAGT | 68 |

[a]Corresponding nucleotide position of GenBank accession number M81413 (PMMoV strain S) [1, 2]

Table S3: qPCR conditions

| Step | Temperature | Time | Cycles |
|---|---|---|---|
| Denaturation | 95 °C | 3 minutes | NA |
| Denaturation | 95 °C | 20 seconds | 40 |
| Annealing | Ta[a] | 1 minute | |
| Extension | 72 °C | 15 seconds | |
| Final extension | 72 °C | 10 minutes | NA |

[a]Ta for PMMoV: 61 °C; Phi6: 63 °C

#### 4 Variability analysis of simplified and rigorous dipstick methods

**(a) Variability chart for mean Ct values for Phi6**

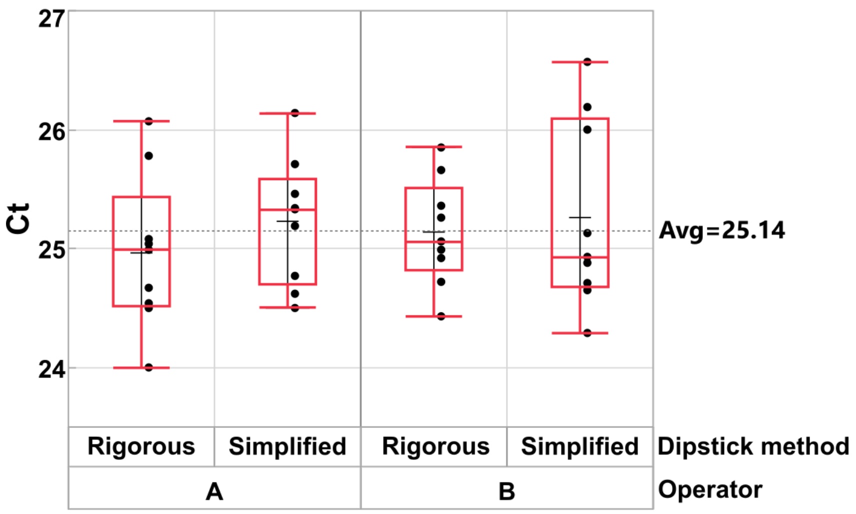


**(b) EMP Gage R&R Results**

| Component | % of Total | 20 40 60 80 |
|---|---|---|
| Gage R&R | 100.0 | |
| Repeatability | 100.0 | |
| Reproducibility | 0.0 | |
| Product Variation | 0.0 | |
| Interaction Variation | 0.0 | |
| Total Variation | 100.0 | |

**(c) Variance Components**

| Component | % of Total | 20 40 60 80 |
|---|---|---|
| Dipstick method | 0.0 | |
| Operator | 0.0 | |
| Dipstick method*Operator | 0.0 | |
| Within | 100.0 | |
| Total | 100.0 | |

Figure. S3: Results obtained for bacteriophage Phi6 RNA captured using rigorous and simplified dipstick methods for sample ID 1: (a) Box plot of Ct values; (b) EMP gage R&R results; and (c) Variance components.

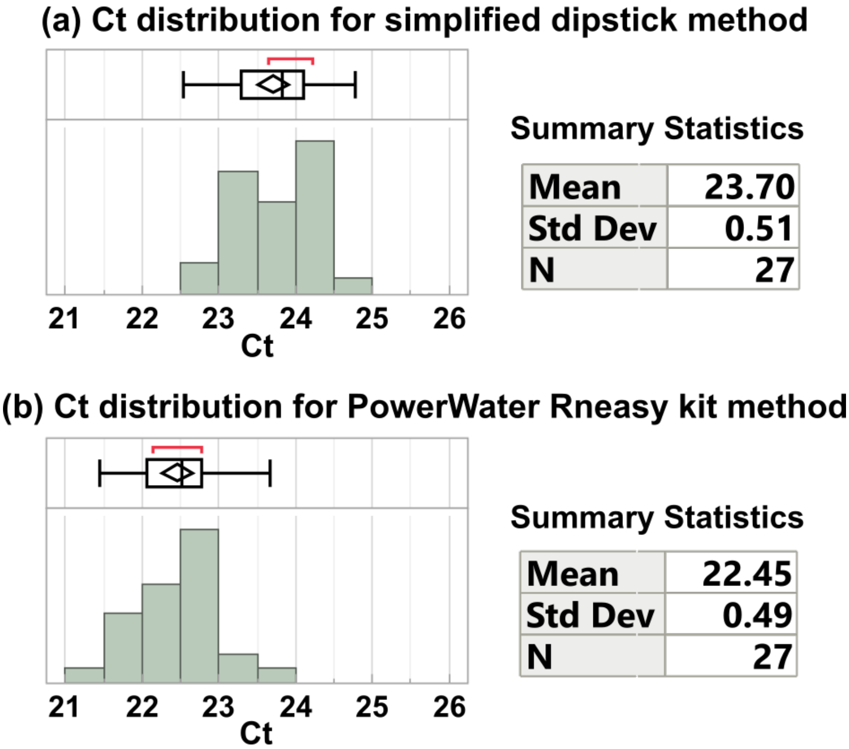


Figure. S4: Ct distribution plot and summary statistics for bacteriophage Phi6 RNA captured from sample IDs 2, 3 and 4 using: (a) simplified dipstick method, and (b) Qiagen PowerWater® RNeasy® kit. Sample ID 5 had more variation compared to other samples. For analysis that included all samples (sample IDs 2 to 5), the mean and standard deviation of Ct obtained with Qiagen PowerWater® RNeasy® kit were 22.41 and 0.47, respectively, whereas the mean and standard deviation of Ct obtained with simplified dipstick method were 23.77 and 0.65, respectively.

### References

[1] Tao Zhang, Mya Breitbart, Wah Heng Lee, Jin-Quan Run, Chia Lin Wei, Shirlena Wee Ling Soh, Martin L Hibberd, Edison T Liu, Forest Rohwer, and Yijun Ruan. Rna viral community in human feces: prevalence of plant pathogenic viruses. *PLoS biology*, 4(1):e3, 2006.

[2] Audrey Garcia, Tri Le, Paul Jankowski, Kadir Yanaç, Qiuyan Yuan, and Miguel Uyaguari-Díaz. Quantitation of human enteric viruses as alternative indicators of fecal pollution to evaluate wastewater treatment processes. *bioRxiv*, pages 2021–08, 2021.
